# Agreement of abdominal palpation with point-of-care ultrasound for hepatosplenic organomegaly: a cross-sectional, population-based study in schistosomiasis-endemic settings of rural Uganda

**DOI:** 10.64898/2026.10.30.26366914

**Authors:** Phyllis Munyiva Isaiah, Eloise S. Ockenden, Christin Puthur, Betty Nabatte, John Bosco Oryema, Hilda Kyarisiima, Prudence Beinamaryo, Victor Anguajibi, Simon Mpooya, Doreen Musiime, Andrew Nuwamanya, Diana Kyomuhangi, Pearl Kobusingye, Euodia Atuhaire, Grace Kabonero, Narcis B. Kabatereine, Goylette F. Chami

## Abstract

**Background:** Ultrasound is commonly used to assess hepatosplenic morbidity in schistosomiasis, but access may be limited in resource-constrained settings. Abdominal palpation is simple and inexpensive, yet its clinical value and diagnostic performance for detecting ultrasound-defined organ enlargement remains uncertain. We evaluated standardised abdominal palpation against point-of-care ultrasound in a schistosomiasis-endemic setting in rural Uganda.

**Methods:** We conducted a cross-sectional study in January-February 2024 of 3177 participants aged 5-92 years from Pakwach, Buliisa and Mayuge districts in the SchistoTrack cohort. Trained nurses who also had midwifery skills performed standardised liver and spleen palpation, assessing palpability, extension, consistency and tenderness. Palpation agreement was assessed against point-of-care B-mode ultrasound performed by sonographers with organ dimensions classified using height-standardised internal reference values. We estimated sensitivity, specificity, predictive values and Cohen’s κ for palpation against ultrasound-defined liver and spleen enlargement and assessed associations between liver consistency and tenderness, and schistosomal periportal fibrosis examined from ultrasound.

**Results:** Spleen palpation had high specificity (98.4%), with sensitivity ranging from 10.9% for moderate splenomegaly to 48.6% for severe splenomegaly. There was strong agreement with ultrasound for severe splenomegaly (κ=0.57) though weak agreement for moderate splenomegaly (κ=0.14). Liver palpation had high specificity (97.7%) but very low sensitivity (3.4-7.3%) and minimal agreement with any hepatomegaly found from ultrasound (κ=0.02-0.04). Among participants with a palpable liver, schistosomal periportal fibrosis was more frequent in those with firm/hard than soft liver consistency (53.3%, n=24/45 vs 8.6%, n=3/35; χ²=17.64, p<0.001); while tenderness was not associated with schistosomal periportal fibrosis. Nurses additionally identified clinically relevant findings, including umbilical collaterals, abdominal swelling, and abdominal or hypochondrial pain and tenderness.

**Conclusions:** Standardised abdominal palpation cannot replace ultrasound for comprehensive detection or exclusion of hepatosplenomegaly. Nevertheless, severe ultrasound-defined splenomegaly was more readily detected by palpation, while liver consistency and other clinical findings provided information beyond organomegaly. In schistosomiasis-endemic settings with limited access to ultrasound, further research is needed to assess building capacity for abdominal palpation and how it can be combined with clinical information to increase clinical suspicion while possibly informing low-cost strategies for triage and referral.

## INTRODUCTION

Hepatosplenic schistosomiasis (HSS) is a chronic manifestation of current or previous infection with *Schistosoma* species that inhabit blood vessels around the intestines (1). HSS can cause hepatic fibrosis and portal hypertension, leading to splenomegaly, ascites, portosystemic collaterals and potentially life-threatening variceal bleeding (2, 3). A rough estimate of 200,000 deaths annually in sub-Saharan Africa have been attributed to these complications (3), however, the true burden may be higher because disability and mortality data from the region remain limited (4). Detecting hepatosplenic morbidity is important in schistosomiasis-endemic settings. Yet, currently there are no World Health Organization (WHO) guidelines for surveying schistosomiasis-related morbidity, and no clinical guidelines exist in most endemic countries for use in primary healthcare facilities.

Point-of-care B-mode ultrasound is the primary technique for the diagnosis of HSS (5), and can identify organ lesions, periportal fibrosis (PPF) patterns, hepatosplenomegaly, abnormal liver texture and ascites (6). However, access to ultrasound machines and the expertise to scan individuals for hepatosplenic abnormalities remains extremely limited in rural, resource-constrained sub-Saharan African settings (7, 8). Physical examination through abdominal palpation is inexpensive and requires minimal clinical expertise. Palpation can provide information on organ palpability, extension, consistency and tenderness (9). However, clinical definitions of hepatosplenomegaly vary considerably (10). Hepatomegaly has been defined as liver extension of >5 cm below the costal margin along the sternal line or >4 cm along the midclavicular line (11), while other definitions use >2 cm (12) below the costal margin or simply the presence of a palpable liver (13). Other approaches consider characteristics such as liver consistency, nodularity and left-lobe prominence (14). Similarly, splenomegaly has been defined either by any palpable spleen or by extension to or beyond the costal margin (15, 16). This variation in definitions, together with differences in examination technique, examiner experience and patient characteristics, leave open questions as to the accuracy and reliability of palpation and its use when ultrasound is unavailable (10, 17–19) and the need for studies to standardise both the palpation technique and criteria used to define hepatosplenomegaly. Further, studies of standardised palpation are limited, missing evaluation against ultrasound measurements adjusted for body height using reference values derived from healthy participants from the same population under study (9, 20, 21).

Therefore, we conducted this study to evaluate the clinical utility of nurse-performed physical examination for assessing hepatosplenic involvement in schistosomiasis across three endemic districts in rural Uganda in 2024. We compared physical examination findings with ultrasound to: (a) determine the diagnostic performance of palpation for detecting ultrasound-defined liver and spleen enlargement; (b) assess associations between liver palpation characteristics and periportal fibrosis (PPF); and (c) determine whether physical examination could identify other clinically important features of hepatosplenic disease beyond organ enlargement.

## METHODS

### Study context

This study was undertaken within the SchistoTrack Cohort, a prospective, community-based cohort established in rural Uganda to investigate schistosome transmission and associated morbidity across Pakwach, Buliisa, and Mayuge districts. Further details of the cohort have been described elsewhere (22).

This was a cross-sectional study conducted in January-February 2024 among 3177 participants aged 5-92 years from Pakwach, Buliisa, and Mayuge districts in rural Uganda. *Schistosoma mansoni,* which is associated with hepatosplenic disease in sub-Saharan Africa, is highly prevalent in the study districts, ranging from 22.05 - 46.64% in 2024 (21). In 2024, schistosomal periportal fibrosis was detected in 18.51% of cohort participants, with a higher prevalence among adults (27.14%) than children (8.53%). Malaria prevalence was estimated at 46.04% in children and 14.12% in adults (21), while Human Immunodeficiency Virus (HIV) and Hepatitis B virus (HBV) occurred at slightly lower prevalence in adults (HIV: 1.19% in children and 6.94% in adults; HBV: 1.49% and 8.46%, respectively) (23), in 2024.

### Physical examination

The standardised palpation protocol used in SchistoTrack was adapted from Vennervald et al. (24) as follows: Four nurses with qualifications in midwifery and experience conducting abdominal palpations conducted the clinical examinations. Each participant was palpated by one nurse. The examination involved initial observation of the abdomen, followed by superficial and deep palpation. Superficial palpation was performed gently, starting in the right lower abdominal quadrant and moving towards the left in a circular motion beneath the ribcage. This initial assessment was used to identify possible organomegaly and tenderness, although findings were recorded only during deep palpation.

For the liver, deep palpation was performed in the right upper abdominal quadrant, with examiners applying pressure to a depth of at least 1 cm unless the liver was markedly enlarged. Tenderness was assessed by observing the participant’s facial response during palpation rather than directly asking about pain. Liver consistency was classified as soft, firm or hard using standardised tactile references corresponding to the lips, nose cartilage and forehead bone, respectively. Extension of the left and right liver lobes below the ribcage was measured separately from the mid-sternal line.

For the spleen, palpation was performed from the left lower abdominal quadrant. Examiners recorded whether the spleen was palpable and assessed tenderness using the same approach as for the liver, applying pressure to a depth of at least 1 cm and observing the participant’s facial response. Splenic extension below the ribcage was measured separately along the left mid-clavicular and mid-axillary lines.

Organ extension was measured in centimetres using a flexible measuring tape, with examiners instructed to look for approximately 2 cm of extension beyond the lower ribcage. A value of 0 cm was recorded when no extension could be measured or the organ was not palpable. In addition to liver and spleen palpation, nurses assessed other clinical findings, including suspected ascites, umbilical collaterals, fever, abdominal swelling, and abdominal or hypochondrial pain and tenderness. Examination findings were recorded electronically using a standardised SchistoTrack nurse palpation survey. A second trained nurse who was working at the same site could be consulted when findings were uncertain. Anthropometric measurements including weight and height also were recorded by nurses.

### Ultrasound assessment

Point-of-care B-mode ultrasound examinations were performed by sonographers trained in the WHO Niamey Protocol using a Philips C5-2 curvilinear transducer and a standardised SchistoTrack protocol. Standardised liver and spleen sweeps were obtained to assess liver patterns, periportal fibrosis (PPF), organ dimensions, ascites and portosystemic collaterals, including splenic, gastro-oesophageal and pancreaticoduodenal varices and splenorenal shunts. PPF was assessed using the WHO Niamey classification of ultrasound liver patterns (5), with patterns C-F classified as indicative of periportal fibrosis.

The left and right liver lobes were measured separately; the left lobe was measured in a parasternal view with the aorta visualised, while the right-lobe measurement incorporated estimation of the upper liver border behind the ribs, with the gallbladder fissure used to delineate the lobes. Spleen length was measured pole-to-pole through the splenic hilum. Before field data collection, sonographers were trained to achieve inter-sonographer agreement within 2 cm for organ-size measurements.

Organ dimensions were classified using previously published height-standardised internal reference values derived from a healthy reference population within the SchistoTrack Cohort (21). The reference population included only participants who were negative for *S. mansoni*, malaria, HBV and HIV, were not pregnant, reported no alcohol consumption within the previous 12 months, and had no ultrasound-detected liver fibrosis. Participants were grouped into height deciles, and organ measurements were standardised against the mean and standard deviation (SD) within the corresponding height decile. For the left and right liver lobes and spleen, measurements were classified as normal, moderately enlarged (≥1 to <2 SD above the reference mean), or severely enlarged (≥2 SD above the reference mean).

### Statistical analysis

Diagnostic performance of abdominal palpation was assessed using 2×2 contingency tables, with ultrasound as the reference standard. Sensitivity, specificity, positive predictive value (PPV) and negative predictive value (NPV) (25) were calculated for splenomegaly and hepatomegaly of the left and right liver lobes. Ultrasound-defined splenomegaly and hepatomegaly were classified as moderate or severe enlargement, with either category considered positive for any organomegaly. Diagnostic performance was also assessed separately for moderate and severe splenomegaly and hepatomegaly, with severe cases excluded from the moderate-enlargement analysis and moderate cases excluded from the severe-enlargement analysis. Overall hepatomegaly was defined as moderate or severe enlargement of either the left or right liver lobe.

Analyses were additionally stratified by age (<15 years and ≥15 years), and sex. The age categories were selected to reflect differences in organ size and palpability related to growth and development in children compared with adults (20). Unlike in adults, for whom normal organ size ranges are well established (20) assessing organ enlargement in children can be more challenging. For example, the spleen may be palpable in up to 10% of healthy children under 15 years of age, yet in most children, it must enlarge to two to three times its normal size before becoming palpable (20).

Confidence intervals for sensitivity, specificity, PPV and NPV were calculated using Wilson’s method, which provides improved interval estimates for proportions, particularly with small samples and proportions close to 0 or 1 (26). Agreement between nurse palpation and ultrasound-defined organ enlargement was assessed using unweighted Cohen’s κ with 95% confidence intervals (27).

Associations between clinical examination and ultrasound findings, as defined in the physical examination and ultrasound assessment sections, were assessed using 2 × 2 contingency tables. Among participants with a palpable liver, associations between ultrasound-defined periportal fibrosis (PPF) and liver consistency (firm/hard versus soft) and liver tenderness (present versus absent) were examined separately. Associations between liver and spleen palpability and between ultrasound-defined liver and spleen enlargement were also assessed.

Fisher’s exact test was used where cell counts were small, and Pearson’s chi-square test was used where appropriate.

## RESULTS

A total of 3222 out of 3385 participants had complete data. Of these, 45 were excluded because abdominal palpation could not be performed due to pain (28), a muscular/bone-like abdomen which could not be palpated (10), or other reasons (7), leaving 3177 participants for analysis (Figure 1).

**Figure 1:**
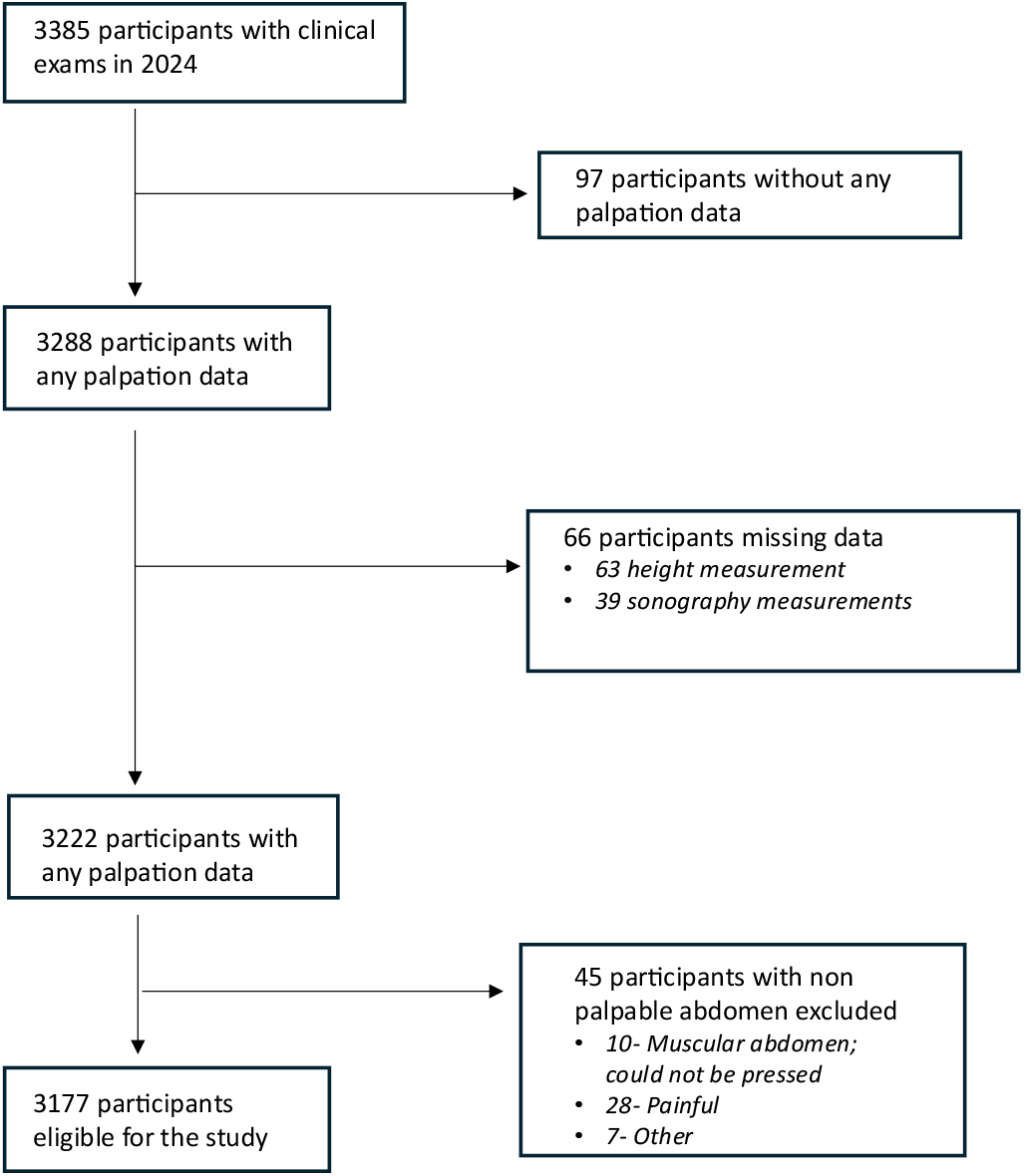
Participant selection flow and study population

### Study population characteristics

Participants ranged in age from 5 to 92 years, with a mean age of 27.0 years (SD 18.6). Of the 3177 participants, 40.3% (n=1279) were children aged <15 years and 59.7% (n=1898) were aged ≥15 years. Overall, 55.3% (n=1756) were female and 44.7% (n=1421) were male. Participants were recruited from Pakwach (43.6%, n=1384), Buliisa (33.0%, n=1049) and Mayuge (23.4%, n=744). The proportion of female participants varied across districts, being highest in Mayuge (63.6%, 473/744), followed by Buliisa (56.9%, 597/1049) and Pakwach (49.6%, 686/1384).

### Splenomegaly

Overall, 8.1% of participants (256/3177) had a palpable spleen. Among participants with a palpable spleen, 43.8% (112/256) had a firm spleen, 24.6% (63/256) had a hard spleen, and 31.6% (81/256) had a soft spleen. Median spleen extension was 3.0 cm (interquartile range (IQR) 0 - 5.0) along the left axillary line and 6.0 cm (IQR 4.0 - 8.0) along the left clavicular line. Spleen palpability was more frequent among children than adults (9.6% vs 7.0%; χ² = 6.68, p=0.010). Among participants with a palpable spleen, adults more frequently had firm or hard spleens and had greater spleen extension than children (all p<0.001; Supplementary Table S1). Ultrasound-defined splenomegaly was present in 26.8% (n=852) of participants, including 16.4% (n=521) with moderate and 10.4% (n=331) with severe splenomegaly. The overall prevalence of splenomegaly was higher among children than adults (30.3%, n=387; vs 24.5%, n=465). Moderate splenomegaly was more common in children (19.5%, n=250) than adults (14.3%, n=271). Among participants with ultrasound-defined splenomegaly, severe splenomegaly accounted for a greater proportion of cases in adults than children (41.7% vs 35.4%; Supplementary Table S2).

### Splenomegaly agreement

Mean ultrasound spleen length was greater among participants with a palpable than non-palpable spleen (15.5; SD 4.0 vs 11.0; SD 2.3 cm). Moderate or severe splenomegaly was observed in 85.2% (218/256) of palpable spleens compared with 21.7% (634/2921) of non-palpable spleens (Table 1). Among participants with a palpable spleen, moderate splenomegaly was more frequent in children than adults (31.7% vs 13.5%), whereas severe splenomegaly was more frequent in adults than children (75.2% vs 49.6%) (Supplementary Table S2). Adults also had greater spleen extension on clinical examination than children (Supplementary Table S1).

**Table 1:** Ultrasound versus nurse palpation findings for splenomegaly.

| ***Ultrasound findings*** | ***Spleen not palpable***<br>***N = 2921*** | ***Spleen palpable***<br>***N = 256*** |
|---|---|---|
| Spleen length, mean (SD), cm | 11.0 (2.3) | 15.5 (4.0) |
| **Height-standardised splenomegaly, n (%)** |  |  |
| Normal spleen length | 2287 (78.3%) | 38 (14.8%) |
| Moderate splenomegaly | 464 (15.9%) | 57 (22.3%) |
| Severe splenomegaly | 170 (5.8%) | 161 (62.9%) |

Table 2 presents the diagnostic performance and agreement of spleen palpation with ultrasound-defined splenomegaly, according to the severity of enlargement. Spleen palpation showed consistently high specificity (98.4%) but varying sensitivity according to the degree of ultrasound-defined splenomegaly. Sensitivity was lowest for moderate splenomegaly (10.9%), increased to 48.6% for severe splenomegaly, and was 25.6% when moderate and severe splenomegaly were combined. Agreement with ultrasound was also greater for severe splenomegaly (κ=0.57, 95% CI 0.52 - 0.62) than for moderate splenomegaly (κ=0.14, 95% CI 0.10 - 0.17).

**Table 2:**
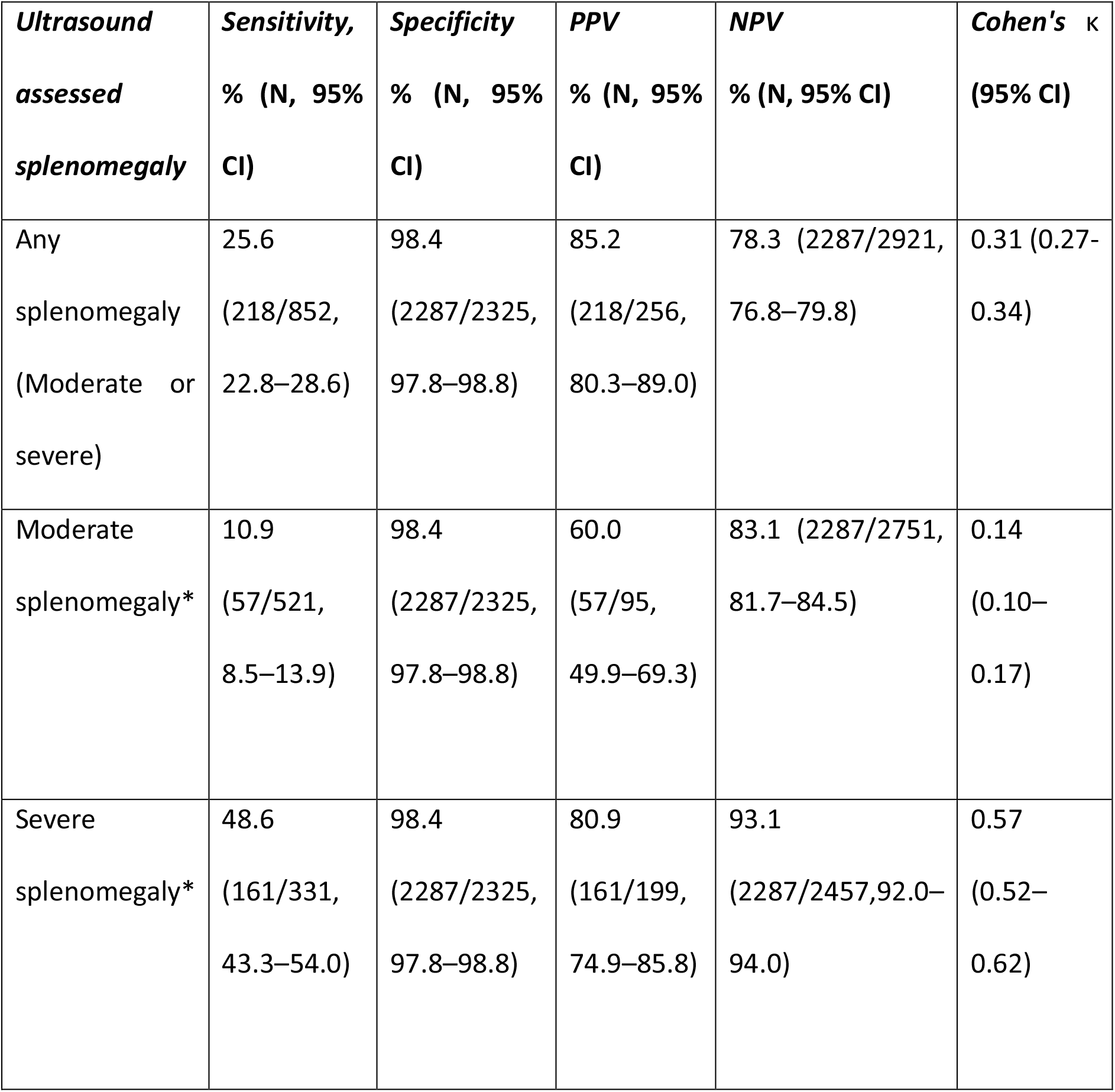
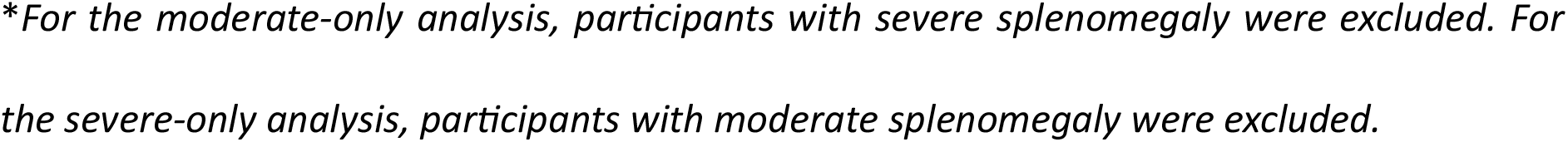
Agreement of abdominal palpation with ultrasound-defined splenomegaly.

Spleen palpation showed high specificity across both age groups (97.4 - 99.0%), while sensitivity was substantially higher for severe splenomegaly in both children (44.5%) and adults (51.5%) than in moderate splenomegaly (15.6% and 6.6%, respectively) (Supplementary Table S3).

### Overlap of portal hypertension signs on ultrasound and splenomegaly

Portosystemic collaterals were identified in 36 participants overall, of whom 63.9% (23/36) had a palpable spleen. Among participants with both a palpable spleen and ultrasound-assessed portosystemic collaterals, 91.3% (21/23) of participants had severe splenomegaly, while one participant had moderate splenomegaly and one participant had no splenomegaly. Among the 852 participants with ultrasound-defined splenomegaly, 3.6% (31/852) had portosystemic collaterals, which were more frequently observed in adults than children (6.0% (28/465) vs 0.8% (3/387); p<0.001, Fisher’s exact test).

Ascites was uncommon (6 participants), with five cases occurring among participants with a palpable spleen.

Nurses independently identified umbilical collaterals in four participants. Of the six participants with ultrasound-detected ascites, two were correctly identified clinically by nurses, while one of 36 participants with ultrasound-detected portosystemic collaterals had umbilical collaterals recorded by nurses. Additional clinical findings recorded by nurses included fever (3.3%; 104/3177) and abdominal swelling (1.4%; 43/3177). Other potentially relevant abdominal findings, including abdominal pain or tenderness, hypochondrial tenderness, and tenderness around the spleen, were recorded in 0.5% (16/3177) of participants (Supplementary Table S4).

### Hepatomegaly

Overall, 2.5% of participants (80/3177) had a palpable liver. Among these participants, 48.8% (39/80) had a firm liver, 7.5% (6/80) a hard liver and 43.8% (35/80) a soft liver, while 65.0% (52/80) had liver tenderness. Liver palpability was similar in children and adults (2.6% vs 2.5%; χ²=0.005, p>0.05), whereas liver consistency differed by age, (Fisher’s exact test, p<0.001), as did liver tenderness (χ²=8.03, p=0.005) (Supplementary Table S5). Among participants with a palpable liver, median left-lobe extension was 6.0 cm (IQR 4.8 - 8.0), while median right-lobe extension was 0 cm (IQR 0 - 3.0).

Ultrasound-defined hepatomegaly was present in 25.9% (824/3177) of participants, involving at least one liver lobe. Left-lobe hepatomegaly was more common, affecting 18.0% (571/3177) of participants, compared with 13.3% (422/3177) with right-lobe hepatomegaly.

Moderate hepatomegaly was detected in 13.9% (443/3177) of left lobes and 12.0% (381/3177) of right lobes, while severe hepatomegaly was present in 4.0% (128/3177) and 1.3% (41/3177), respectively.

Mean ultrasound-measured liver lobe dimensions were slightly greater among participants with a palpable liver than those without a palpable liver, for both the right lobe (9.5 vs 9.1 cm) and left lobe (8.6 vs 8.1 cm). Moderate or severe hepatomegaly was present in 21.3% (17/80) of right lobes and 26.3% (21/80) of left lobes among participants with a palpable liver, compared with 13.1% (405/3097) and 17.7% (550/3097), respectively, among those without a palpable liver (Table 3).

**Table 3:** Ultrasound versus nurse palpation findings for hepatomegaly.

| **Ultrasound findings** | ***Liver not palpable***<br>N = 3097 | ***Liver palpable***<br>N =80 |
|---|---|---|
| ***Right liver lobe*** |  |  |
| Right liver lobe, mean (SD), cm | 9.1 (1.9) | 9.5 (1.8) |
| Normal right liver lobe | 2692 (86.9%) | 63 (78.8%) |
| Moderate hepatomegaly of the right liver lobe | 367 (11.9%) | 14 (17.5%) |
| Severe hepatomegaly of the right liver lobe | 38 (1.2%) | 3 (3.8%) |
| ***Left liver lobe*** |  |  |
| Left liver lobe, mean (SD), cm | 8.1 (1.6) | 8.6 (1.8) |
| Normal left liver lobe | 2547 (82.2%) | 59 (73.8%) |
| Moderate hepatomegaly of the left liver lobe | 428 (13.8%) | 15 (18.8%) |
| Severe hepatomegaly of the left liver lobe | 122 (3.9%) | 6 (7.5%) |

Among participants with a palpable liver, right-lobe enlargement was more frequent in children than adults (30.3% vs 14.9%), whereas left-lobe enlargement was more frequent in adults than children (34.0% vs 15.1%) (Supplementary Table S6).

### Hepatomegaly agreement

Liver palpation had low sensitivity (3.4 - 7.3%) but high specificity (97.7%) for ultrasound-defined hepatomegaly, indicating that most ultrasound-detected hepatomegaly was not identified by palpation. Agreement between palpation and ultrasound was minimal (κ=0.02 - 0.04) (Table 4). Age-stratified analyses similarly showed low sensitivity and high specificity of liver palpation for ultrasound-defined hepatomegaly Sensitivity for any left-lobe enlargement was 2.6% in children and 4.2% in adults, while sensitivity for any right-lobe enlargement was 7.6% and 2.4%, respectively (Supplementary Table S7). Estimates for severe right-lobe enlargement were imprecise because of the small number of cases, particularly among children (Supplementary Table S7).

**Table 4:** Agreement of nurse palpation with ultrasound-defined hepatomegaly.

| **Ultrasound defined hepatomegaly** | **Sensitivity, % (N, 95% CI)** | **Specificity, % (N, 95% CI)** | **PPV, % (N, 95% CI)** | **NPV, % (N, 95% CI)** | ***Cohen's*** **κ (95% CI)** |
|---|---|---|---|---|---|
| **Left liver lobe** |  |  |  |  |  |
| Any hepatomegaly (moderate or severe) | 3.7 (21/571, 2.4-5.6) | 97.7 (2547/2606, 97.1-98.2) | 26.2 (21/80, 17.9-36.8) | 82.2 (2547/3097, 80.9-83.5) | 0.02 (-0.003-0.05) |
| Moderate hepatomegaly* | 3.4 (15/443, 2.1-5.5) | 97.7 (2547/2606, 97.1-98.2) | 20.3 (15/74, 12.7-30.8) | 85.6 (2547/2975, 84.3-86.8) | 0.02 (-0.01-0.04) |
| Severe hepatomegaly* | 4.7 (6/128, 2.2-9.8) | 97.7 (2547/2606, 97.1-98.2) | 9.2 (6/65, 4.3-18.7) | 95.4 (2547/2669, 94.6-96.2) | 0.03 (-0.02-0.08) |
| **Right liver lobe** |  |  |  |  |  |
| Any hepatomegaly (moderate or severe) | 4.0 (17/422, 2.5-6.4) | 97.7 (2692/2755, 97.1-98.2) | 21.2 (17/80,13.7-31.4) | 86.9 (2692/3097, 85.7-88.1) | 0.03 (-0.003-0.06) |
| Moderate hepatomegaly* | 3.7 (14/381, (2.2-6.1) | 97.7 (2692/2755, 97.1-98.2) | 18.2 (14/77, 11.2-28.2) | 88.0 (2692/3059, 86.8-89.1) | 0.02 (-0.01-0.05) |
| Severe hepatomegaly* | 7.3 (3/41, 2.5-19.4) | 97.7 (2692/2755, 97.1-98.2) | 4.5 (3/66, 1.6-12.5) | 98.6 (2692/2730, 98.1-99.0) | 0.04 (-0.02-0.10) |

\**For moderate and severe hepatomegaly analyses, severe and moderate hepatomegaly cases were excluded from the denominator rather than treated as normal respectively.*

### Association between liver consistency/ tenderness with schistosomal PPF

Among participants with a palpable liver (n=80), schistosomal PPF was more frequent in participants with a firm or hard liver than in participants with a soft liver (53.3%, 24/45 vs 8.6%, 3/35; χ²=17.64, p<0.001). In contrast, PPF was present in 36.5% (19/52) of participants with liver tenderness compared with 28.6% (8/28) of participants without tenderness, with no significant association between liver tenderness and PPF (χ²=0.52, p>0.05).

### Association between liver and spleen palpability and ultrasound-defined enlargement

Liver and spleen palpability frequently co-occurred, with 51.3% (41/80) of participants with a palpable liver also having a palpable spleen (χ²=206.65, p<0.001; Supplementary Table S8). Similarly, among participants with ultrasound-defined hepatomegaly, 35.1% (289/824) also had ultrasound-defined splenomegaly (χ²=38.07, p<0.001).

## DISCUSSION

There is a need to identify complementary strategies to ultrasound in areas that lack such equipment. Abdominal palpation is commonly applied in resource poor settings with several highly trained staff due to the availability of nurses and midwives for prenatal care. Here we assessed the agreement of abdominal palpation with point-of-care B-mode ultrasound for hepatosplenomegaly across 3177 individuals from communities endemic with intestinal schistosomiasis in rural Uganda. The diagnostic performance of standardised abdominal palpation varied by organ and severity. Spleen palpation was substantially more sensitive for severe than moderate splenomegaly, whereas liver palpation had low sensitivity for both liver lobes. Despite this limited diagnostic performance, clinical examination provided information beyond organ size; firm or hard liver consistency was positively associated with schistosomal PPF, while umbilical collaterals and other clinically relevant findings also were identified.

Spleen palpation showed consistently high specificity (98.4%) but limited sensitivity, which varied by the severity of ultrasound-defined splenomegaly. Sensitivity was higher for severe (48.6%) than moderate splenomegaly (10.9%), with a similar pattern observed in children and adults. The limited sensitivity observed in our study is broadly consistent with previous studies showing that physical examination may fail to detect ultrasound-defined splenomegaly (28–30). The limited diagnostic performance may partly reflect variation in how splenomegaly is defined by palpation and ultrasound. In our study, splenic extension below the costal margin was measured in centimetres, and nearly all participants with a palpable spleen (95.3%, 244/256) had an extension of at least 2 cm at one or both measurement lines. We therefore used palpability as a simple binary clinical indicator rather than applying a specific extension threshold in the diagnostic-performance analyses. In contrast, ultrasound-defined splenomegaly was determined using spleen length relative to height-standardised reference values derived from a healthy population within the same cohort (21).

Hackett’s classification, one of the most widely used clinical grading systems for palpable splenomegaly in field surveys (31, 32), is an ordinal system that incorporates palpability and the position of the lowest palpable point relative to anatomical landmarks, including the costal margin, umbilicus and pubic symphysis. Although this provides a standardised approach to describing palpable splenomegaly, Hackett grades do not necessarily correspond directly to ultrasound-defined spleen size (33). The importance of how palpable splenomegaly is defined has been demonstrated previously by Gerspacher-Lara et al. (15) in an *S. mansoni*-endemic population. Considering any palpable spleen as positive yielded 72.2% sensitivity and 90.5% specificity for ultrasound-defined splenomegaly, whereas requiring >4 cm extension below the costal margin reduced sensitivity to 27.8% but increased specificity to 98.0% (15). These findings show that the clinical threshold used to define splenomegaly influences diagnostic performance. There is a need to understand whether any palpable spleen is sufficient to raise clinical suspicion. Although, previous studies have shown that clinically palpable spleens are not necessarily enlarged or pathological (34) . As such, additional standardised studies focusing on measuring extension below the costal margin as done here are needed to identify relevant cut-offs that best reflect long-term patient outcomes focused on the study or clinical context. Focusing on patient outcomes is important as there may also be discrepancies from ultrasound due to probe positioning during examination, variations in spleen position and shape, respiratory movements and acoustic shadowing from ribs and bowel gas (35, 36).

Although spleen palpation had low sensitivity overall in the present study, its high specificity may help identify individuals with more pronounced splenic abnormalities who could be prioritised for further assessment. This prioritisation is particularly relevant to hepatosplenic schistosomiasis, where severe splenomegaly may signal more advanced disease and warrant assessment for portal-hypertension-related complications, including portosystemic collaterals and gastro-oesophageal varices, which can be life-threatening (1, 37, 38). In our study, of the 36 participants with ultrasound-detected portosystemic collaterals, 23 had a palpable spleen, of whom 21 had severe splenomegaly. These findings suggest that spleen palpation may have some value for clinical case identification and referral including capturing other underlying pathologies, rather than as a stand-alone method for screening or excluding splenomegaly.

Liver palpation had low sensitivity (3.4-7.3%) though high specificity (97.7%) for ultrasound-assessed hepatomegaly. These findings are consistent with previous studies showing that liver palpability does not reliably reflect ultrasound-assessed hepatomegaly (7,30). Several factors may contribute to the low sensitivity of liver palpation. Although a problem with other studies (7) due to inconsistently defined reference populations, SchistoTrack used an internally defined healthy reference population to assess the hepatomegaly via ultrasound (21). Liver consistency may influence palpation, especially since a recently enlarged liver tends to be of soft consistency and more difficult to palpate than a firm or hard liver (39). In cases of marked enlargement, the liver may extend into the lower abdomen, making its lower border difficult to identify if palpation does not begin sufficiently low (39). Conversely, deep inspiration may bring a normal-sized liver below the costal margin, making it palpable (40). Anatomical variations, such as Riedel’s lobe, where part of the right liver lobe extends below the costal margin, may also cause a normal-sized liver to be palpable and mistaken for an enlarged liver (41). Taken together, these factors may contribute to both missed cases of hepatomegaly and the incorrect identification of normal-sized livers as enlarged.

Age-stratified analyses showed a similar overall pattern in children and adults, with spleen palpation more sensitive for severe than moderate splenomegaly and liver palpation having low sensitivity for both children and adults. The greater proportion of moderate splenomegaly among children reported in this study may partly reflect malaria exposure, which is also known to exacerbate *S. mansoni*-associated splenomegaly in children (42). Given that malaria is routinely treated in the study area (43), including through SchistoTrack, treatment could limit malaria-associated splenomegaly and may partly explain the lower frequency of severe splenomegaly among children. Age-related anatomical differences may also influence spleen palpability, as a palpable spleen can occur in otherwise healthy children and does not necessarily indicate pathological enlargement (44). In adults, severe splenomegaly may reflect longer-term exposure to *S. mansoni* alongside other causes of splenic enlargement, including HIV (21) and malaria (45). Though contextually interesting, the relative contribution of these infections to splenic enlargement was beyond the scope of this agreement analysis and already studied thoroughly elsewhere for ultrasound (21).

Although palpation had low sensitivity for detecting ultrasound-defined liver and spleen enlargement, it may still provide important information about the presence and severity of hepatosplenic morbidity. In particular, severe splenomegaly is more likely to be detected by palpation and is an important clinical feature of advanced hepatosplenic schistosomiasis. Liver consistency may also provide useful information. Among participants with a palpable liver, schistosomal PPF was present in 53.3% of individuals with a firm or hard liver compared with 8.6% of those with a soft liver. Evidence from other *S. mansoni*-endemic settings has been mixed. In Brazil, clinical findings including a hard or nodular liver were associated with ultrasound-detected schistosomal PPF (10), while in Venezuela, hard liver consistency was associated with *S. mansoni* infection (46). In contrast, Marinho et al. reported moderate agreement between hardened liver consistency and ultrasound-detected fibrosis in Brazil (30). These findings suggest that liver consistency may provide information about underlying schistosomal PPF, but further studies are needed to establish its reliability as a clinical marker. Clinical examination may also identify other features of advanced disease. In addition to severe splenomegaly and changes in liver consistency, examination in our study identified umbilical collaterals, which may occur with portal hypertension (1, 47); as well as fever, abdominal swelling, and abdominal or hypochondrial pain and tenderness. These findings highlight the complementary value of bedside examination in capturing clinical information beyond ultrasound-defined hepatosplenic abnormalities. This finding is consistent with evidence from other clinical settings showing that physical examination provides immediate, low-cost clinical information and, despite the limited sensitivity of some individual findings, can increase clinical suspicion and guide further diagnostic investigation (48–50). In schistosomiasis-endemic settings with limited access to imaging, combining palpation findings, other clinical signs and presenting symptoms may therefore help identify individuals who warrant further assessment or referral. Future studies should evaluate whether integrating these features into a standardised clinical assessment algorithm improves case identification and prioritisation for further investigation.

There were several limitations to this study. Both palpation and ultrasound are examiner-dependent and may be influenced by examination technique and participant characteristics. Although sonographers were trained to achieve agreement within 2 cm for organ-size measurements, comparable inter-examiner reproducibility was not formally assessed for palpation, despite the use of a standardised protocol. Nurses also identified cases of suspected ascites that were not confirmed by ultrasound. Given the high sensitivity of ultrasound for detecting free abdominal fluid, these findings were likely to represent false-positive clinical assessments rather than ascites missed on ultrasound. This may partly reflect a lack of standardisation in the use of palpation and percussion for assessing ascites. Other causes of abdominal distension, including adiposity, flatulence, and pregnancy, may also have been mistaken for ascites. Further, only 80 participants had a palpable liver, limiting analyses on consistency or clinical findings.

## CONCLUSIONS

Overall, palpation cannot replace point-of-care ultrasound or reliably exclude hepatosplenic morbidity but may increase clinical suspicion, supporting case identification and referral by identifying individuals who warrant further assessment where access to ultrasound is limited. Bedside assessment also captures clinical history and presenting symptoms, including fever, abdominal pain and abdominal swelling, which, when considered alongside physical examination findings, may strengthen clinical suspicion and guide further investigation. If validated in other settings, our findings might begin to provide a basis for developing and validating a simple, low-cost clinical assessment algorithm that combines palpation findings, clinical signs and presenting symptoms to identify individuals who should be prioritised for further investigation or referral for ultrasound or other investigations in schistosomiasis-endemic settings.

## Data Availability

All data generated or analysed during this study are included in this published article and its supplementary information files.

## LIST OF ABBREVIATIONS

CI: Confidence interval
HBV: Hepatitis B virus
HIV: Human Immunodeficiency Virus
HSS: Hepatosplenic Schistosomiasis
IQR: Interquartile Range
NPV: Negative Predictive Value
PPF: Periportal Fibrosis
PPV: Positive Predictive Value
*S.*: *mansoni Schistosoma mansoni*
SD: Standard Deviation
WHO: World Health Organisation

## DECLARATIONS

### Ethics approval and consent to participate

This study was approved by Oxford Tropical Research Ethics Committee (OxTREC 509-21), Vector Control Division Research Ethics Committee of the Uganda Ministry of Health (VCDREC146), and Uganda National Council for Science and Technology (UNCST HS 1664ES). Written Informed consent was obtained from all study participants or their legal guardians before their involvement. Participants were informed of their right to decline participation or withdraw from the study at any stage without consequences. Strict confidentiality was maintained throughout the research process to protect participants’ personal information.

### Consent for publication

Not applicable.

### Author contributions

Conceptualization: GFC. Data curation: GFC, PMI, EO, CP, BN, JBO, VA, SM, MD, NA, DK, KP, AE, GK, NBK. Formal analysis: PMI, EO, CP. Investigation: PMI, BN, VA, SM, MD, NA, DK, KP, AE, GK.

Methodology, visualization: PMI, GFC. Writing – original draft: PMI, GFC. Validation: PMI, GFC. Writing – review and editing: GFC, PMI, EO, CP, BN, JBO, HK, PB, VA, SM, MD, NA, DK, KP, AE, GK, NBK. Funding acquisition and supervision: GFC. Resources: GFC.

All authors read and approved the final manuscript.

## Acknowledgements

We are grateful to our study participants, and the SchistoTrack teams, especially the nurses and sonographers. We also like to thank the Uganda Ministry of Health, and the local district leaders. Special thanks also to the Oxford team for everyday discussions, and feedback.

