## Supplementary Material for "Agreement of abdominal palpation with point-of-care ultrasound for hepatosplenic organomegaly: a cross-sectional, population-based study in schistosomiasis-endemic settings of rural Uganda"

**Supplementary Tables S1 – S8.**

**S1. Spleen palpability**

| Characteristic | <15 years | ≥15 years | Test statistic* | P-value |
|---|---|---|---|---|
| **Spleen palpability (N=3177)** | | | $\chi^2$ = 6.68 | 0.010 |
| Yes | 123 (9.6%) | 133 (7.0%) | | |
| No | 1156 (90.4%) | 1765 (93.0%) | | |
| **Spleen consistency (n=256)** | | | $\chi^2$ = 50.71 | <0.001 |
| Firm | 41 (33.3%) | 71 (53.4%) | | |
| Hard | 17 (13.8%) | 46 (34.6%) | | |
| Soft | 65 (52.8%) | 16 (12.0%) | | |
| **Spleen extension (cm), median (IQR)** | | | | |
| Spleen extension, left axillary line (cm) | 2.0 (0.0, 4.0) | 4.0 (1.0, 7.0) | W= 5462 | <0.001 |
| Spleen extension, left clavicular line (cm) | 5.0 (4.0, 7.0) | 8.0 (6.0, 10.0) | W = 4309 | <0.001 |

*Pearson's chi-square test was used for categorical variables, and the Wilcoxon rank-sum test was used to compare spleen extension between children and adults.

**S2. Ultrasound versus nurse palpation findings for splenomegaly**

| ***Ultrasound findings*** | ***Adults (≥15 years) Spleen not palpable N = 1765*** | ***Adults (≥15 years) Spleen palpable N = 133*** | ***Children (<15 years) Spleen not palpable N = 1156*** | ***Children (<15 years) Spleen palpable N = 123*** |
|---|---|---|---|---|
| Normal spleen length | 1418 (80.3%) | 15 (11.3%) | 869 (75.2%) | 23 (18.7%) |
| Moderate splenomegaly | 253 (14.3%) | 18 (13.5%) | 211 (18.3%) | 39 (31.7%) |
| Severe splenomegaly | 94 (5.3%) | 100 (75.2%) | 76 (6.6%) | 61 (49.6%) |

**S3. Agreement of nurse palpation with ultrasound-defined splenomegaly by age and sex**

| **Spleen any enlargement** | **Sensitivity % (N) [95% CI]** | **Specificity % (N) [95% CI]** | **PPV % (N) [95% CI]** | **NPV % (N) [95% CI]** | **Cohen's κ (95% CI)** |
|---|---|---|---|---|---|
| Whole population | 25.6 (218/852) [22.8-28.6] | 98.4 (2287/2325) [97.8-98.8] | 85.2 (218/256) [80.3-89.0] | 78.3 (2287/2921) [76.8-79.8] | 0.31 (0.27-0.34) |
| Children (<15 years) | 25.8 (100/387) [21.7-30.4] | 97.4 (869/892) [96.2-98.3] | 81.3 (100/123) [73.5-87.2] | 75.2 (869/1156) [72.6-77.6] | 0.29 (0.24-0.34) |

| | | | | | |
|---|---|---|---|---|---|
| Adults (>=15 years) | 25.4 (118/465) [21.6-29.5] | 99.0 (1418/1433) [98.3-99.4] | 88.7 (118/133) [82.2-93.0] | 80.3 (1418/1765) [78.4-82.1] | 0.32 (0.27-0.37) |
| Female | 24.3 (103/423) [20.5-28.7] | 98.6 (1314/1333) [97.8-99.1] | 84.4 (103/122) [77.0-89.8] | 80.4 (1314/1634) [78.4-82.3] | 0.30 (0.25-0.35) |
| Male | 26.8 (115/429) [22.8-31.2] | 98.1 (973/992) [97.0-98.8] | 85.8 (115/134) [78.9-90.7] | 75.6 (973/1287) [73.2-77.9] | 0.31 (0.26-0.36) |
| **Moderate splenomegaly** | | | | | |
| Whole population | 10.9 (57/521) [8.5-13.9] | 98.4 (2287/2325) [97.8-98.8] | 60.0 (57/95) [49.9-69.3] | 83.1 (2287/2751) [81.7-84.5] | 0.14 (0.10-0.17) |
| Children (<15 years) | 15.6 (39/250) [11.6-20.6] | 97.4 (869/892) [96.2-98.3] | 62.9 (39/62) [50.5-73.8] | 80.5 (869/1080) [78.0-82.7] | 0.18 (0.12-0.24) |
| Adults (>=15 years) | 6.6 (18/271) [4.2-10.3] | 99.0 (1418/1433) [98.3-99.4] | 54.5 (18/33) [38.0-70.2] | 84.9 (1418/1671) [83.1-86.5] | 0.09 (0.04-0.13) |
| Female | 8.8 (23/260) [6.0-12.9] | 98.6 (1314/1333) [97.8-99.1] | 54.8 (23/42) [39.9-68.8] | 84.7 (1314/1551) [82.8-86.4] | 0.11 (0.06-0.16) |
| Male | 13.0 (34/261) [9.5-17.7] | 98.1 (973/992) [97.0-98.8] | 64.2 (34/53) [50.7-75.7] | 81.1 (973/1200) [78.8-83.2] | 0.16 (0.10-0.21) |
| **Severe splenomegaly** | | | | | |
| Whole population | 48.6 (161/331) [43.3-54.0] | 98.4 (2287/2325) [97.8-98.8] | 80.9 (161/199) [74.9-85.8] | 93.1 (2287/2457) [92.0-94.0] | 0.57 (0.52-0.62) |
| Children (<15 years) | 44.5 (61/137) [36.5-52.9] | 97.4 (869/892) [96.2-98.3] | 72.6 (61/84) [62.3-81.0] | 92.0 (869/945) [90.0-93.5] | 0.50 (0.42-0.59) |
| Adults (>=15 years) | 51.5 (100/194) [44.6-58.5] | 99.0 (1418/1433) [98.3-99.4] | 87.0 (100/115) [79.6-91.9] | 93.8 (1418/1512) [92.5-94.9] | 0.61 (0.55-0.68) |
| Female | 49.1 (80/163) [41.5-56.7] | 98.6 (1314/1333) [97.8-99.1] | 80.8 (80/99) [72.0-87.4] | 94.1 (1314/1397) [92.7-95.2] | 0.58 (0.50-0.65) |
| Male | 48.2 (81/168) [40.8-55.7] | 98.1 (973/992) [97.0-98.8] | 81.0 (81/100) [72.2-87.5] | 91.8 (973/1060) [90.0-93.3] | 0.56 (0.48-0.63) |

**S4. Clinically important findings by palpation (N= 3177)**

| Palpation clinical findings | **n (%)** |
|---|---|
| Febrile patient | 104 (3.3%) |
| Abdominal swelling | 43 (1.4%) |
| Other potentially relevant abdominal findings* | 16 (0.5%) |
| Scars | 15 (0.5%) |
| Umbilical collaterals | 4 (0.1%) |

**S5. Liver palpability**

| **Characteristic** | **<15 years** | **≥15 years** | **Test** | **P-value** |
|---|---|---|---|---|
| **Liver palpability (N=3177)** | | | $\chi^2$=0.005 | 0.946 |
| Yes | 33 (2.6%) | 47 (2.5%) | | |
| No | 1246 (97.4%) | 1851 (97.5%) | | |
| **Liver consistency (n=80)** | | | Fisher's exact | <0.001 |
| Firm | 7 (21.2%) | 32 (68.1%) | | |
| Hard | 1 (3.0%) | 5 (10.6%) | | |
| Soft | 25 (75.8%) | 10 (21.3%) | | |
| **Liver tenderness (n=80)** | | | $\chi^2$=8.03 | 0.005 |
| No | 18 (54.5%) | 10 (21.3%) | | |
| Yes | 15 (45.5%) | 37 (78.7%) | | |

Pearson's chi-square test was used to compare liver palpability and tenderness between children and adults, while Fisher's exact test was used to compare liver consistency because of small cell counts.

**S6. Ultrasound versus nurse palpation findings for hepatomegaly**

| ***Ultrasound findings*** | ***Adults (≥15 years) Liver not palpable (N=1851)*** | ***Adults (≥15 years) Liver palpable (N=47)*** | ***Children (<15 years) Liver not palpable (N=1246)*** | ***Children (<15 years) Liver palpable (N=33)*** |
|---|---|---|---|---|
| **Right liver lobe** | | | | |
| Normal right liver lobe | 1568 (84.7%) | 40 (85.1%) | 1124 (90.2%) | 23 (69.7%) |
| Moderate hepatomegaly of the right liver lobe | 251 (13.6%) | 6 (12.8%) | 116 (9.3%) | 8 (24.2%) |
| Severe hepatomegaly of the right liver lobe | 32 (1.7%) | 1 (2.1%) | 6 (0.5%) | 2 (6.1%) |
| **Left liver lobe** | | | | |
| Normal left liver lobe | 1490 (80.5%) | 31 (66.0%) | 1057 (84.8%) | 28 (84.8%) |

| | | | | |
|---|---|---|---|---|
| Moderate hepatomegaly of the left liver lobe | 272 (14.7%) | 11 (23.4%) | 156 (12.5%) | 4 (12.1%) |
| Severe hepatomegaly of the left liver lobe | 89 (4.8%) | 5 (10.6%) | 33 (2.6%) | 1 (3.0%) |

**S7. Agreement of nurse palpation with ultrasound-defined hepatomegaly by age and sex**

| **Ultrasound finding** | **Sensitivity % (N) [95% CI]** | **Specificity % (N) [95% CI]** | **PPV % (N) [95% CI]** | **NPV % (N) [95% CI]** | **Cohen's κ (95% CI)** |
|---|---|---|---|---|---|
| **LEFT LIVER LOBE -** Any enlargement | | | | | |
| Whole population | 3.7 (21/571) [2.4-5.6] | 97.7 (2547/2606) [97.1-98.2] | 26.2 (21/80) [17.9-36.8] | 82.2 (2547/3097) [80.9-83.5] | 0.02 (-0.003-0.05) |
| Children (<15 years) | 2.6 (5/194) [1.1-5.9] | 97.4 (1057/1085) [96.3-98.2] | 15.2 (5/33) [6.7-30.9] | 84.8 (1057/1246) [82.7-86.7] | 0.00 (-0.04-0.04) |
| Adults (>=15 years) | 4.2 (16/377) [2.6-6.8] | 98.0 (1490/1521) [97.1-98.6] | 34.0 (16/47) [22.2-48.3] | 80.5 (1490/1851) [78.6-82.2] | 0.03 (0.001-0.07) |
| Female | 2.0 (7/349) [1.0-4.1] | 98.6 (1387/1407) [97.8-99.1] | 25.9 (7/27) [13.2-44.7] | 80.2 (1387/1729) [78.3-82.0] | 0.01 (-0.02-0.03) |
| Male | 6.3 (14/222) [3.8-10.3] | 96.7 (1160/1199) [95.6-97.6] | 26.4 (14/53) [16.4-39.6] | 84.8 (1160/1368) [82.8-86.6] | 0.04 (-0.004-0.09) |
| **Moderate hepatomegaly** | | | | | |
| Whole population | 3.4 (15/443) [2.1-5.5] | 97.7 (2547/2606) [97.1-98.2] | 20.3 (15/74) [12.7-30.8] | 85.6 (2547/2975) [84.3-86.8] | 0.02 (-0.01-0.04) |
| Children (<15 years) | 2.5 (4/160) [1.0-6.3] | 97.4 (1057/1085) [96.3-98.2] | 12.5 (4/32) [5.0-28.1] | 87.1 (1057/1213) [85.1-88.9] | 0.00 (-0.04-0.04) |
| Adults (>=15 years) | 3.9 (11/283) [2.2-6.8] | 98.0 (1490/1521) [97.1-98.6] | 26.2 (11/42) [15.3-41.1] | 84.6 (1490/1762) [82.8-86.2] | 0.03 (-0.01-0.06) |
| Female | 1.9 (5/267) [0.8-4.3] | 98.6 (1387/1407) [97.8-99.1] | 20.0 (5/25) [8.9-39.1] | 84.1 (1387/1649) [82.3-85.8] | 0.01 (-0.02-0.04) |
| Male | 5.7 (10/176) [3.1-10.1] | 96.7 (1160/1199) [95.6-97.6] | 20.4 (10/49) | 87.5 (1160/1326) [85.6-89.2] | 0.04 (-0.02-0.09) |

| | | | | | |
|---|---|---|---|---|---|
| | | | [11.5-33.6] | | |
| **Severe hepatomegaly** | | | | | |
| Whole population | 4.7 (6/128) [2.2-9.8] | 97.7 (2547/2606) [97.1-98.2] | 9.2 (6/65) [4.3-18.7] | 95.4 (2547/2669) [94.6-96.2] | 0.03 (-0.02-0.08) |
| Children (<15 years) | 2.9 (1/34) [0.5-14.9] | 97.4 (1057/1085) [96.3-98.2] | 3.4 (1/29) [0.6-17.2] | 97.0 (1057/1090) [95.8-97.8] | 0.00 (-0.06-0.07) |
| Adults (>=15 years) | 5.3 (5/94) [2.3-11.9] | 98.0 (1490/1521) [97.1-98.6] | 13.9 (5/36) [6.1-28.7] | 94.4 (1490/1579) [93.1-95.4] | 0.05 (-0.02-0.11) |
| Female | 2.4 (2/82) [0.7-8.5] | 98.6 (1387/1407) [97.8-99.1] | 9.1 (2/22) [2.5-27.8] | 94.5 (1387/1467) [93.3-95.6] | 0.02 (-0.04-0.07) |
| Male | 8.7 (4/46) [3.4-20.3] | 96.7 (1160/1199) [95.6-97.6] | 9.3 (4/43) [3.7-21.6] | 96.5 (1160/1202) [95.3-97.4] | 0.06 (-0.03-0.14) |
| **RIGHT LIVER LOBE - Any enlargement** | | | | | |
| Whole population | 4.0 (17/422) [2.5-6.4] | 97.7 (2692/2755) [97.1-98.2] | 21.2 (17/80) [13.7-31.4] | 86.9 (2692/3097) [85.7-88.1] | 0.03 (-0.003-0.06) |
| Children (<15 years) | 7.6 (10/132) [4.2-13.4] | 98.0 (1124/1147) [97.0-98.7] | 30.3 (10/33) [17.4-47.3] | 90.2 (1124/1246) [88.4-91.7] | 0.08 (0.02-0.15) |
| Adults (>=15 years) | 2.4 (7/290) [1.2-4.9] | 97.5 (1568/1608) [96.6-98.2] | 14.9 (7/47) [7.4-27.7] | 84.7 (1568/1851) [83.0-86.3] | 0.00 (-0.03-0.03) |
| Female | 2.8 (8/282) [1.4-5.5] | 98.7 (1455/1474) [98.0-99.2] | 29.6 (8/27) [15.9-48.5] | 84.2 (1455/1729) [82.4-85.8] | 0.02 (-0.01-0.06) |
| Male | 6.4 (9/140) [3.4-11.8] | 96.6 (1237/1281) [95.4-97.4] | 17.0 (9/53) [9.2-29.2] | 90.4 (1237/1368) [88.7-91.9] | 0.04 (-0.02-0.10) |
| **Moderate hepatomegaly** | | | | | |
| Whole population | 3.7 (14/381) [2.2-6.1] | 97.7 (2692/2755) [97.1-98.2] | 18.2 (14/77) [11.2-28.2] | 88.0 (2692/3059) [86.8-89.1] | 0.02 (-0.01-0.05) |
| Children (<15 years) | 6.5 (8/124) [3.3-12.2] | 98.0 (1124/1147) [97.0-98.7] | 25.8 (8/31) [13.7-43.2] | 90.6 (1124/1240) [88.9-92.1] | 0.07 (0.002-0.13) |
| Adults (>=15 years) | 2.3 (6/257) [1.1-5.0] | 97.5 (1568/1608) [96.6-98.2] | 13.0 (6/46) [6.1-25.7] | 86.2 (1568/1819) [84.5-87.7] | 0.00 (-0.03-0.03) |

| | | | | | |
|---|---|---|---|---|---|
| Female | 2.8 (7/252) [1.4-5.6] | 98.7 (1455/1474) [98.0-99.2] | 26.9 (7/26) [13.7-46.1] | 85.6 (1455/1700) [83.8-87.2] | 0.02 (-0.01-0.06) |
| Male | 5.4 (7/129) [2.7-10.8] | 96.6 (1237/1281) [95.4-97.4] | 13.7 (7/51) [6.8-25.7] | 91.0 (1237/1359) [89.4-92.4] | 0.03 (-0.03-0.08) |
| **Severe hepatomegaly** | | | | | |
| Whole population | 7.3 (3/41) [2.5-19.4] | 97.7 (2692/2755) [97.1-98.2] | 4.5 (3/66) [1.6-12.5] | 98.6 (2692/2730) [98.1-99.0] | 0.04 (-0.02-0.10) |
| Children (<15 years) | 25.0 (2/8) [7.1-59.1] | 98.0 (1124/1147) [97.0-98.7] | 8.0 (2/25) [2.2-25.0] | 99.5 (1124/1130) [98.8-99.8] | 0.11 (-0.04-0.26) |
| Adults (>=15 years) | 3.0 (1/33) [0.5-15.3] | 97.5 (1568/1608) [96.6-98.2] | 2.4 (1/41) [0.4-12.6] | 98.0 (1568/1600) [97.2-98.6] | 0.00 (-0.05-0.06) |
| Female | 3.3 (1/30) [0.6-16.7] | 98.7 (1455/1474) [98.0-99.2] | 5.0 (1/20) [0.9-23.6] | 98.0 (1455/1484) [97.2-98.6] | 0.02 (-0.05-0.10) |
| Male | 18.2 (2/11) [5.1-47.7] | 96.6 (1237/1281) [95.4-97.4] | 4.3 (2/46) [1.2-14.5] | 99.3 (1237/1246) [98.6-99.6] | 0.06 (-0.03-0.15) |

**S8. Association between nurse-detected liver and spleen palpability**

| **Nurse palpation finding** | **Spleen not palpable** | **Spleen palpable** | **Total** | $\chi^2$* | **p-value** |
|---|---|---|---|---|---|
| Liver not palpable | 2882 (93.1%) | 215 (6.9%) | 3097 | | |
| Liver palpable | 39 (48.8%) | 41 (51.3%) | 80 | | |
| **Total** | **2921 (91.9%)** | **256 (8.1%)** | **3177** | 206.65 | <0.001 |

*Pearson's chi-square test used
